# Impact of CMV Infection and Cancer Treatment on Vaccine Efficacy in Oncology Patients

**DOI:** 10.1101/2025.08.21.25334174

**Authors:** Darshak K. Bhatt, Manas Joshi, Frederique Visscher, Annemarie Boerma, Sjoukje F Oosting, Astrid A M van der Veldt, T Jeroen N Hiltermann, Corine H Geurts van Kessel, Anne-Marie C Dingemans, Egbert F Smit, John B A G Haanen, Elisabeth G E de Vries, Debbie van Baarle

## Abstract

Cytomegalovirus (CMV) infection leaves lasting effects on the immune system, particularly shaping T-cell populations. In patients with advanced cancer, persistent CMV exposure may influence vaccine responses due to immunosuppression from disease progression or prior therapies. To explore this, we developed *CMVision*, an open-source ELISpot scoring pipeline that goes beyond traditional spot-forming unit counts, enabling accurate analysis of complex T-cell responses, even in wells with overlapping or ambiguous spots. Using *CMVision*, we assessed SARS-CoV-2 spike-specific responses after mRNA vaccination in patients with cancer with receiving chemotherapy, immunotherapy or both combined and in cancer-free controls from the VOICE study. CMV-specific responses remained stable across groups. Notably, individuals with stronger CMV-reactivity showed enhanced spike-specific responses, suggesting CMV-memory may reflect immunological “fitness.” These findings position *CMVision* as a valuable tool and highlight CMV-reactivity as a potential biomarker for vaccine readiness in cancer patients.

## Introduction

Chronic viral exposures, such as cytomegalovirus (CMV), may influence how the human immune system responds to infections and vaccination^1,2^. CMV is a ubiquitous beta-herpesvirus that establishes lifelong latency and leads to persistent immune activation and remodeling of the T-cell pool of an individual^1^. In particular, CMV drives the accumulation of differentiated, oligoclonal memory T-cell subsets, simultaneously influencing the overall T-cell pool^3^. While typically asymptomatic in immunocompetent individuals, CMV’s immunological imprint becomes increasingly relevant in older adults^2,4^ and immunocompromised populations^5–7^, such as patients with advanced cancers undergoing chemotherapy or immunotherapy. Despite growing awareness of CMV’s influence on immune homeostasis, its role in modulating immune responses to novel antigens, such as those introduced via vaccination, remains less understood.

Biologically, the effect of CMV on immune fitness and vaccine responsiveness is nuanced and context-dependent. Multiple studies have demonstrated that CMV alters the composition and function of the T-cell compartment, frequently inducing terminal differentiation and the expansion of effector memory T-cells re-expressing CD45RA (EMRA)^8–11^. In the elderly, these changes have been associated with reduced responsiveness to vaccines, particularly those targeting the influenza virus^12^. However, the literature presents a complex picture: CMV has been implicated in both enhancing and suppressing immune responses. Some studies report that CMV infection can potentiate responses to heterologous antigens, possibly via bystander activation or stimulation of the innate immune system^8^, while others describe impaired memory CD4 T-cell responses to unrelated pathogens^9^.

A limited but growing number of studies have explored CMV’s impact on immune responses to SARS-CoV-2 vaccination. Notably, CMV seropositivity has been associated with phenotypic shifts toward immune senescence and altered NK and T-cell function^13^; however, it does not appear to impair SARS-CoV-2-specific antibody production or the durability of vaccine-induced memory responses^14^. Another study^15^, however, observed reduced spike-specific T-cell responses to vaccination in CMV-seropositive individuals who had not previously encountered SARS-CoV-2, suggesting that prior SARS-CoV-2 antigenic exposure may mitigate potential CMV-associated immunological constraints. These findings underscore the need to move beyond binary classifications of CMV serostatus and instead consider the magnitude or gradient of CMV-specific T-cell reactivity as a more informative variable. In one of our previous studies, we examined how low, medium, and high levels of CMV-specific T-cell responses corresponded to influenza infection outcomes, but did not observe any significant differences^16^. Although CMV adds significant immunological complexity, few studies have directly examined how differences in CMV-specific reactivity affect vaccine responsiveness, particularly in vulnerable populations such as patients with cancer.

This question is especially timely given the increasing use of mRNA vaccines in immunocompromised populations. Patients with cancer patients, due to therapies and disease-related immunosuppression, often display varied immune responsiveness. Yet, previous work from the VOICE study has shown that exposure to cancer therapy alone does not uniformly impair T-cell responses to SARS-CoV-2 vaccination^17,18^. However, the potential role of chronic viral infection, especially the magnitude of CMV-specific immunity, as a determinant of vaccine responsiveness in this group remains understudied.

Accurately quantifying T-cell responses in such heterogeneous populations requires robust and sensitive analytical tools. The ELISpot assay remains a widely used method for measuring antigen-specific cytokine production at the single-cell level. Conventionally, ELISpot outputs are quantified by counting discrete spot-forming units (SFUs), a practice that captures broad immune activation but can miss subtle qualitative differences in T-cell function. Particularly in samples with overlapping, faint, or morphologically diverse spots (such as those from immunocompromised individuals) this count-based approach may fail to capture the full spectrum of immune responses^19–21^.

Recent computational tools have aimed to improve ELISpot analysis using automated image processing and machine learning^19–21^. However, most approaches remain limited to spot enumeration and do not incorporate additional metrics such as spot intensity, well-level signal, spatial distribution, or occupancy, features that could more accurately reflect biologically relevant T-cell activity. This technical gap is particularly problematic in studies requiring high-resolution discrimination of immune phenotypes, such as investigations into vaccine responsiveness among cancer patients with variable CMV exposure histories. To address these analytical and biological gaps, we developed *CMVision*, a robust, open-source ELISpot scoring pipeline designed to move beyond traditional spot count–based quantification. *CMVision* incorporates multiple metrics, including spot intensity distributions, well-level occupancy, and spatial patterning, enabling more nuanced characterization of immune responses even in visually ambiguous wells. We applied *CMVision* to analyze spike-specific T-cell responses following SARS-CoV-2 mRNA vaccination in participants stratified by cancer therapy treatment from the VOICE study, alongside controls without cancer.

## Methods

### Patient material and ELISpot assay

Peripheral blood mononuclear cell (PBMC) samples were obtained from participants enrolled in the previously published VOICE trial^17,18^, a prospective, multi-center clinical study evaluating mRNA-1273 vaccine responses in patients with cancer and controls ClinicalTrials.gov, NCT04715438. In total, 791 individuals without prior SARS-CoV-2 infection were enrolled across four cohorts: controls without cancer (CTRL, n = 247), patients treated with immune checkpoint inhibitors (IT, n = 137), patients receiving chemotherapy (CT, n = 244), and those treated with a combination of chemotherapy and immunotherapy (CT/IT, n = 163). PBMCs were collected at baseline (prior to vaccination) and at multiple time points following the second vaccine dose (28 days, 6, 11, 12, and 18 months). This study utilized cryopreserved PBMCs from that biorepository. In particular, we analyzed samples from baseline (T0) and 28 days post-vaccination (T2), selecting a subset of participants with high-quality ELISpot images and complete metadata. Cohort demographics are summarized in **Table 1**.

**Table 1:** Demographics and clinical characteristics of patients involved in the study.

| Variable | Statistic | Value |
| --- | --- | --- |
| Age | Mean ± SD | 61.7 ± 11.4 |
|  | Range | 19 – 87 |
| Gender | Female | 234 (48%) |
|  | Male | 254 (52%) |
| Tumor stage | Stage-I | 10 (2%) |
|  | Stage-II | 28 (5.7%) |
|  | Stage-III | 57 (11.7%) |
|  | Stage-IV | 226 (46.3%) |
|  | NA | 167 (34.2%) |
| Cohort | Chemotherapy (CT) | 141 (28.9%) |
|  | Chemotherapy and Immunotherapy (CT/IT) | 94 (19.3%) |
|  | Healthy (CTRL) | 166 (34%) |
|  | Immunotherapy (IT) | 87 (17.8%) |
| VoiceID | Unique Ids | 488 |
| Primary tumor localisation | Bone, Articular cartilage and Soft tissues | 2 (0.4%) |
|  | Breast | 51 (10.5%) |
|  | Central nervous system | 6 (1.2%) |
|  | Digestive tract | 46 (9.4%) |
|  | Endocrine glands | 2 (0.4%) |
|  | Female genital organs | 10 (2%) |
|  | Head and neck | 4 (0.8%) |
|  | Male genital organs | 11 (2.3%) |
|  | Other/unspecified sites | 1 (0.2%) |
|  | Respiratory tract | 117 (24%) |
|  | Skin | 44 (9%) |
|  | Urinary tract | 28 (5.7%) |
|  | NA | 166 (34%) |

ELISpot assays were performed as previously described. Briefly, MultiScreen HTS IP filter plates (Millipore, MSIPS4510) were ethanol-activated and coated overnight at 4°C with anti-human IFNγ antibody (Mabtech, 3420-3-250, 5 μg/mL). After blocking with X-VIVO medium supplemented with 2% human serum for 1 hour at 37°C, thawed PBMCs were incubated in the same medium for 60 minutes. A total of 2×10⁵ PBMCs were stimulated in triplicate for 20-24 hours at 37°C with two separate peptide pools covering the SARS-CoV-2 spike protein and the CMV-pp65 protein (JPT, 0.5 μg/peptide/mL). DMSO and phytohemagglutinin (PHA, 2 μg/mL) served as negative and positive controls, respectively. Plates were developed using biotinylated anti-IFNγ detection antibody (1:1000, diluted, Mabtech, 3420-6-250), streptavidin poly-HRP (Sanquin, M2051), and TMB substrate (Mabtech, 3651-10).

### ELISpot Image Acquisition and Preprocessing

The resulting well images from the ELISpot reader (AID iSpot) were exported and used for this analysis, as illustrated in **Figure 1**. Raw color images were processed using *CellProfiler*^22^ (version 4.2.6; Broad Institute) with a custom image analysis pipeline provided in the Supplementary Material. In brief, images were converted from color to grayscale, and pixel intensities were rescaled to the range of 0 to −1. Wells were identified as primary objects using the three-class Otsu thresholding method with the “assign foreground” option. Within each well, cytokine spots were segmented using a separate two-class Otsu threshold. For each well, multiple quantitative features were extracted, including spot count, mean and integrated intensity values for both spots and the entire well, mean and total spot area, and the proportion of the well area occupied by signal (well occupancy). All processed images and corresponding binary masks were saved, and extracted features were exported to spreadsheets for downstream analysis.

**Figure 1:**
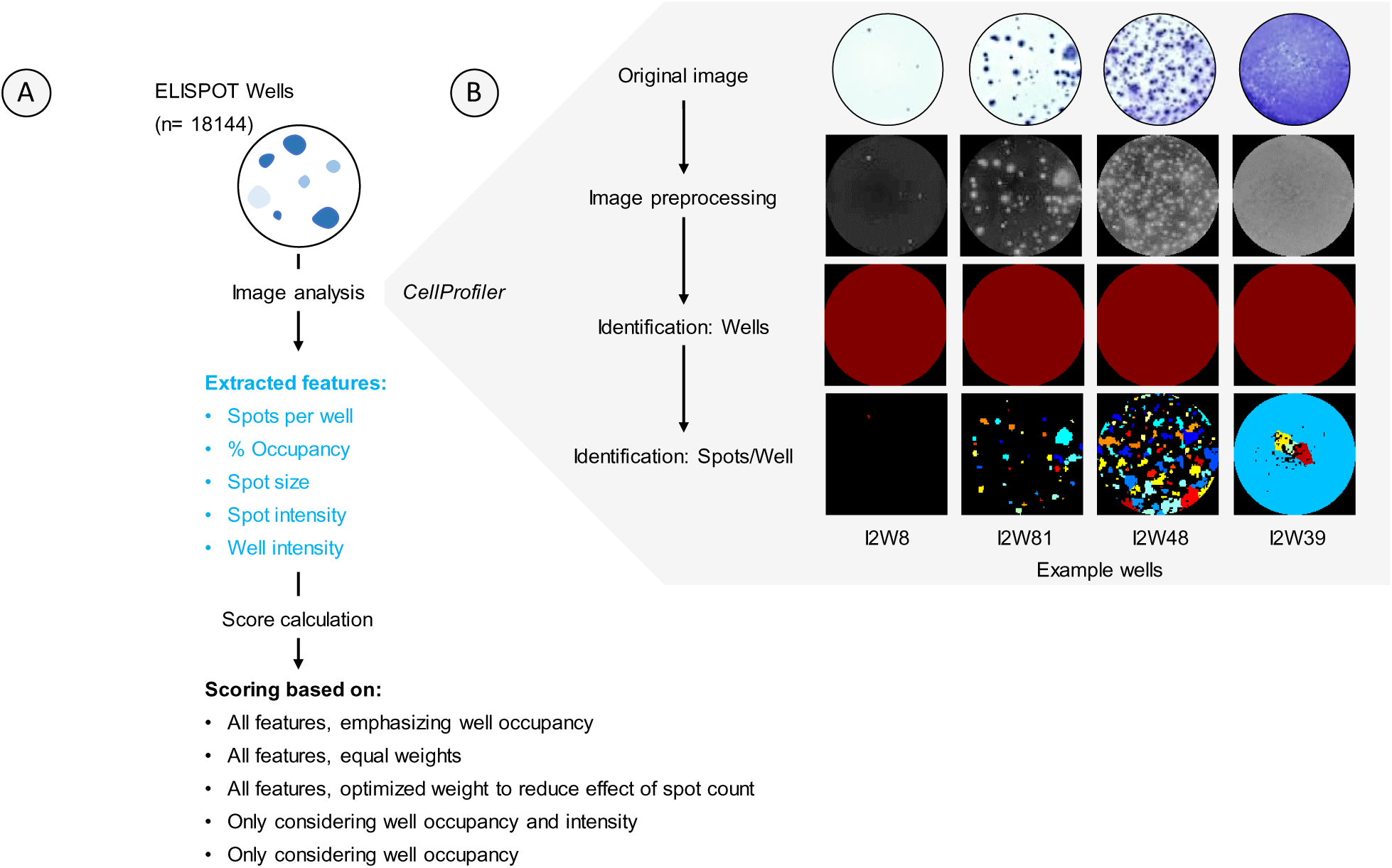
Overview of the CMVision pipeline for ELISpot image analysis and composite scoring. (A) The workflow includes image acquisition, feature extraction, and generation of multiple scoring systems to quantify T-cell responses, with feature extraction processed via CellProfiler. We used 18,144 images, with a well per image, taken from a 96 well plate. From over 100 features, we selected the most biologically meaningful ones: spots per well, % well occupancy (area covered by all spots in a well), spot size, mean spot intensity, and integrated well intensity. These features were further processed to calculate 5 different combined scores, either using all or selected features. (B) Example wells that were processed and analyzed using the pipeline, illustrating the accuracy of identifying respective wells and spots per well for the analysis.

### Feature Analysis and Scoring

To evaluate and compare ELISpot well reactivity across conditions, we developed five scoring systems based on image-derived features extracted from CellProfiler. All subsequent analyses were conducted in R (version 4.5.1)^23^. Six quantitative variables were selected for this purpose: spot count per well (*C*), percent of well area occupied (*O*), mean spot intensity (*I_s_*), mean spot area (*A*), integrated well intensity (*I_w_*), and a background penalty term derived from well-level intensity (*B*). All missing or non-finite values were set to zero to penalize poor-quality wells. A reference “proxy” score was constructed by z-scoring four key features (occupancy, spot intensity, spot area, and background penalty) and averaging them per well. This composite proxy served as the optimization target for evaluating score performance using Spearman correlation.

The first scoring system, *Emphasize Well Occupancy*, is a composite formula that includes all six features and applies tunable weights to key variables and more weight to well occupancy. It is defined as:

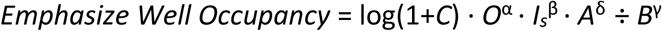

The exponents α, β, γ, and δ were optimized through a grid search to maximize correlation with the proxy score, allowing emphasis to be placed on specific aspects, such as occupancy or intensity, depending on data quality. The *Emphasize Well Occupancy* score was designed to prioritize well occupancy features in particular. During grid search optimization, we intentionally favored parameter combinations that up-weighted occupancy (*O*) and down-weighted background penalty (*B*), while allowing moderate influence from spot-level features such as intensity and area. This score is particularly suited for wells where spatial coverage and total signal footprint are more informative than discrete spot counts, such as in highly reactive or confluent wells.

The second scoring system, *Equal Weights*, retains the same structure but applies equal weighting to all components, thereby offering a balanced metric:

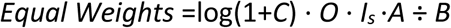

A third system, *Optimized Weights*, was generated by independently re-performing the grid search to find a distinct set of optimal exponents, potentially capturing an alternative balance of signal features.

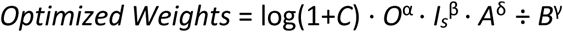

In contrast to the *Emphasize Well Occupancy* score, the *Optimized Weights* score was generated through a separate, fully data-driven optimization. No constraints were placed on which features should be emphasized. Instead, the goal was to maximize Spearman correlation with a composite proxy score constructed from normalized feature values. Interestingly, this unconstrained optimization consistently assigned a low weight to spot count (*C*), suggesting that in our dataset, spot count alone was a weaker predictor of overall well signal compared to occupancy, intensity, or spot morphology. As a result, *Optimized Weights* reflects an empirical model that down-weights spot enumeration in favor of features more aligned with well-level activity.

In addition to these comprehensive scoring methods, we also defined two simplified metrics. The *Well Occupancy and Intensity* score captures broad well-level activation by multiplying occupancy with well intensity. This score omits spot-level features entirely and focuses on total well engagement.

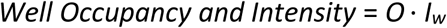

Lastly, the *Well Occupancy* score uses occupancy alone as a minimal, baseline metric. This single-feature score offers interpretability and computational simplicity while reflecting an immunological signal in many contexts.

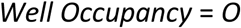

Figure 2 visually compares the scoring outcomes, highlighting how each metric ranks well in reactivity through a heatmap and representative well images.

**Figure 2:**
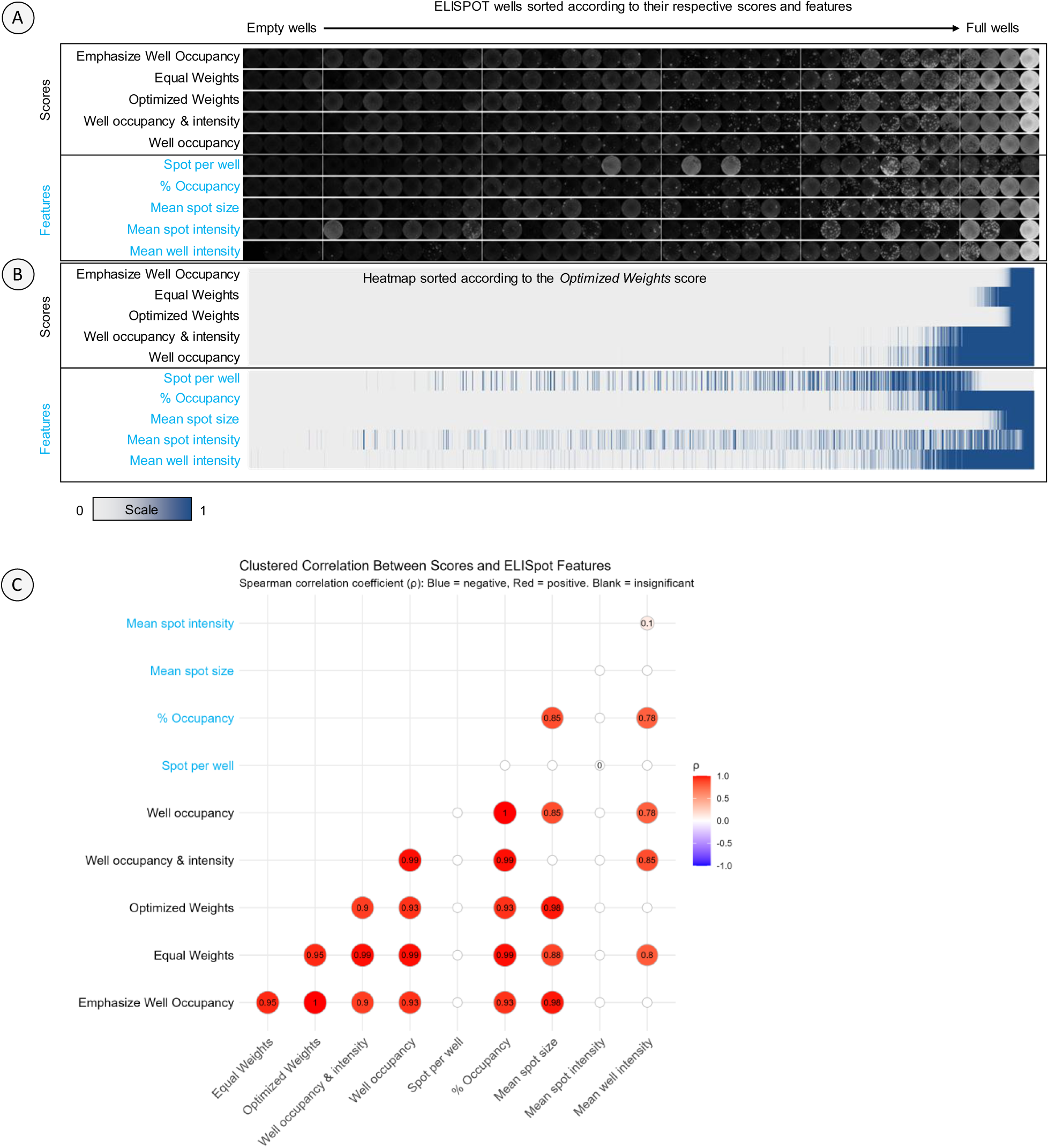
Comparative ranking, distribution, and correlation of ELISpot features and composite scores. **(A)** Representative images from 40 wells, illustrating well morphology and spot distribution across different scores and features, ranked accordingly. **(B)** Heatmap displaying normalized data from 18,144 wells, including features and scores, ordered by the Optimized Weights score. The wells are scored on a scale of 0 (low) to 1 (high) for each respective feature and score. **(C)** Correlation matrix depicting pairwise relationships between ELISpot scores and individual features across all wells. Spearman correlation coefficients are shown to highlight feature interdependencies. Blank entries indicate non-significant correlation (p-value > 0.05).

### Data Aggregation and Normalization

Scores were aggregated at the individual, antigen, and time point levels. For each patient tagged with a unique VoiceID, technical replicates were averaged to generate a single value per condition. DMSO control well scores were subtracted from antigen-stimulated wells to correct for background signal. Spike-specific responses were measured using two overlapping peptide pools, targeting S1 and S2 regions of the protein. These were averaged to yield a single composite spike score per time point, consistent with the well-based scoring approach.

### Statistical Analyses and Plotting

All statistical analyses and visualizations were performed using R. Comparative analyses included changes in spike-specific responses at baseline (T0) and 28 days after vaccination (T2), changes in CMV-specific responses between timepoints, associations between immune response and age, and the relationship between CMV-specific reactivity and changes in spike-specific responses. Paired comparisons were assessed using Wilcoxon signed-rank tests. Correlation analyses were conducted using Spearman’s rank method. All plots and statistical results are presented in the figures, with detailed p-values and correlation coefficients included in the respective figure legends.

### Pipeline and Code Availability

The *CMVision* pipeline and related code generated in this work can be found at https://github.com/d-bhatt/CMVision.

## Results

### Development and validation of the *CMVision* scoring pipeline

We first developed and validated *CMVision*, a computational pipeline for automated ELISpot image analysis and scoring (Figure 1A). The pipeline processed over 18,000 well images obtained from ELISpot assays using PBMCs from cancer patients and healthy controls. Using *CellProfiler*, we extracted multiple biologically meaningful features per well, including spot count, spot size, spot intensity, integrated well intensity, and percentage of well area occupied by signal. These features were combined using multiple scoring systems designed to enhance interpretability and accuracy of immunological quantification, particularly in wells with high spot overlap or variable morphology. The pipeline successfully identified wells and their constituent spots across a wide range of well intensities and morphologies, including low-responding, sparse wells and highly confluent or saturated wells (Figure 1B).

#### Visualization and performance of feature-based scores

To evaluate the discriminative power of the scores and features, we visualized 40 representative wells ranked by each score and individual feature (Figure 2A). We found that all composite scores generally ranked high-responding wells similarly, especially the *Optimized Weights*, *Equal Weights*, and *Emphasize Well Occupancy* scores. The simpler *Well Occupancy* score, however, sometimes failed to prioritize wells with high intensity due to its lack of intensity weighting. In contrast, rankings based on individual features such as spot count or mean spot intensity were inconsistent. For instance, some high-responding wells with dense and bright signals received low rankings under the spot count metric, reflecting how high-density wells can appear under-counted due to overlapping spots.

This finding was supported by a heatmap of 18,144 wells, ranked by the *Optimized Weights* score (Figure 2B). We chose this score because it consistently captured wells with strong biological responses across diverse conditions, balancing multiple features such as intensity, spot count, and occupancy. The heatmap shows strong concordance among the three composite scores, while the individual features displayed variable alignment, with spot count often showing poor association with well reactivity. Percent occupancy and well intensity are strongly associated with composite score ranking.

#### Inter-feature and inter-score correlations

A correlation matrix between all features and composite scores (Figure 2C) revealed strong correlations between the composite scoring systems (Spearman’s ρ > 0.8), indicating that the scores are internally consistent despite differences in construction. Among the individual features, percent occupancy showed the strongest correlation with the composite scores, while spot count and mean spot intensity showed the weakest correlation. These results validate our scoring approach and highlight the limitations of relying solely on spot enumeration in high-throughput T-cell analysis.

#### Spike-specific T-cell responses increase after vaccination across all patient groups

Using the Optimized Weights score, we next evaluated vaccine-induced T-cell responses to SARS-CoV-2 spike antigen across therapy groups by including samples from participants with high-quality ELISpot data and complete metadata, as described in Table 1. Spike-specific responses significantly increased from baseline (T0) to two weeks post-boost (T2) across all patient cohorts (Figure 3A), including those treated with chemotherapy, immunotherapy, or both. The magnitude of response varied slightly across treatment groups, but did not abrogate the vaccine-induced T-cell response, as previously observed in our earlier study^17^. Robust spike-specific responses were observed across all age groups and cohorts, with no significant correlation with age (p-value >0.05). However, in the group receiving immunotherapy, these responses exhibited a weak inverse correlation with age (Figure 3C).

**Figure 3:**
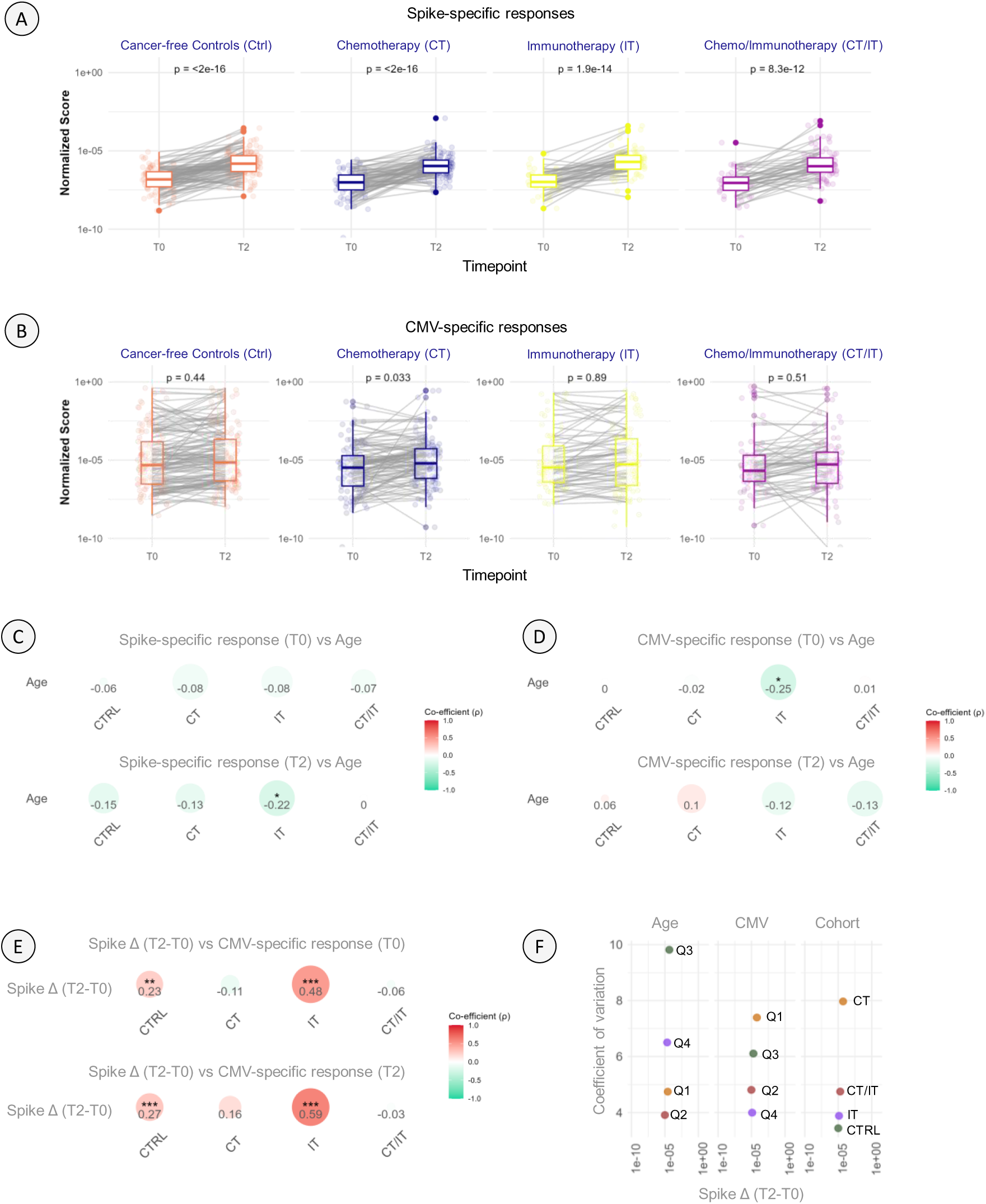
T-cell responses in patients with cancer and controls, controls zijn niet healthy following SARS-CoV-2 mRNA vaccination. (A) Spike-specific T-cell responses stratified by cancer therapy cohorts at baseline (T0) and post-vaccination (T2). (B) CMV-specific T-cell responses stratified by cancer therapy cohorts at T0 and T2. (C) Correlation between spike-specific responses at T0 and T2 and patient age. (D) Correlation between CMV-specific responses at T0 and T2 and patient age. (E) Correlation between spike-specific response change (T2–T0) and CMV-specific responses at T0 and T2. (F) Variation in spike-specific response change (T2–T0) with respect to patient age, CMV-specific response, or therapy cohort. Paired statistical analyses were used to compare responses between T0 and T2. In panels (C–E), correlation coefficients and corresponding p-values are indicated. Statistical significance was defined as p < 0.05 and denoted as follows: * = p < .05, ** = p < .01, *** = p < .001. In panel (F), the coefficient of variation was calculated as the ratio of the standard deviation to the mean. Patients were divided into quartiles (Q1–Q4) based on age or the magnitude of CMV-specific response. Cohort abbreviations based on cancer therapy: CT = chemotherapy, CTRL = control (no cancer), IT = Immunotherapy, CT/IT = Chemotherapy and Immunotherapy.

#### CMV-specific T-cell responses are stable and therapy-independent

Next, we assessed whether CMV-specific T-cell responses changed post-vaccination or differed across cohorts. CMV responses showed no significant changes between T0 and T2 (Figure 3B). In the group receiving chemotherapy, there was a small yet significant increase in the CMV-specific responses post-vaccination, which may reflect low-level activation of CMV memory or chemotherapy-induced immune turnover. Overall, these results suggest the CMV-specific memory T-cell responses remain stable and are not significantly impacted by mRNA vaccination or cancer treatment. Unlike spike responses, CMV reactivity did not show age-dependent changes (Figure 3D), suggesting that CMV memory may be maintained even in older and immunocompromised individuals, as indicated in literature.

#### CMV-specific reactivity correlates with vaccine-induced spike T-cell response

Finally, we tested whether the strength of CMV-specific reactivity correlated with spike-specific vaccine responsiveness, measured as the change in spike-specific responses (T2-T0). As shown in Figure 3D, a significant positive correlation was observed between CMV and spike-specific vaccine responsiveness, particularly in patients treated with immunotherapy. This suggests that individuals with stronger CMV memory may possess a more "fit" T-cell compartment capable of mounting stronger responses to novel antigens. However, this correlation was attenuated in patients who received chemotherapy, with or without immunotherapy, possibly reflecting broader T-cell dysfunction in these groups.

#### Variation in vaccine responses is associated with cancer therapy and CMV-specific reactivity

The coefficient of variation (CV) for change in spike-specific responses (T2–T0) was highest in the cohort receiving chemotherapy or both chemo- and immunotherapy (Figure 3F). This indicated a greater inter-individual heterogeneity in vaccine-induced T-cell expansion among patients receiving chemotherapy. In contrast, only immunotherapy-treated patients and controls displayed lower CV values, suggesting more consistent responses in these groups. When stratified by CMV-specific response quartiles, individuals in the lowest quartile showed greater variability in spike responses, whereas those in the highest CMV quartile exhibited lower variability. This pattern could reflect that individuals capable of mounting strong CMV responses also tend to generate more consistent spike-specific memory responses. Age quartiles showed non-linear and irregular differences in CV compared to therapy or CMV quartiles, indicating that chronological age alone may be less influential on response variability than immune history or treatment exposure in this cohort.

## Discussion

In this study, we developed and applied *CMVision*, a robust, open-source ELISpot image analysis pipeline that integrates multiple image-derived metrics to provide more comprehensive and nuanced quantification of T-cell responses. By combining well occupancy, spot morphology, and intensity features into composite scores, we propose an approach that surpasses traditional spot count–based methods, which often fail in wells with overlapping or faint spots, particularly in immunocompromised samples. Using this pipeline, we analyzed spike-specific and CMV-specific T-cell responses in cancer patients and healthy controls from the VOICE study, offering new insight into how chronic viral memory, particularly CMV, relates to vaccine-induced immunity.

Conventional ELISpot quantification relies heavily on counting discrete spot-forming units, which may underestimate responses in confluent or high-density wells. Our composite scoring approach revealed that spot count often poorly correlates with other biologically meaningful features, such as well occupancy or signal intensity. These results are consistent with recent work emphasizing the need for improved ELISpot quantification tools using machine learning or advanced image analysis^19–21^. However, existing tools generally focus on count optimization and rarely integrate well-level intensity or spatial metrics. *CMVision* addresses this gap, providing a scalable method for multidimensional ELISpot scoring.

Our findings show that mRNA vaccination induces robust spike-specific T-cell responses in patients during various systemic cancer therapy histories, including chemotherapy, immunotherapy, or chemo-immunotherapy. This is consistent with earlier *VOICE* study results using conventional spot counts^17,18^, and extends these findings by demonstrating similar trends using a feature-integrated scoring approach. Importantly, our results support the view that even immunocompromised patients, such as those recently treated with cytotoxic therapies, can mount meaningful cellular response to SARS-CoV-2 vaccination. Age correlated inversely but weakly and often non-significantly with spike-specific responses, aligning with reports of immunosenescence in elderly individuals^8,9^.

Overall, the CMV-specific T-cell responses remained remarkably stable over time and across therapy groups, except for patients treated with chemotherapy, where a slight increase was observed. This aligns with previous observations that CMV memory is long-lived and largely unaffected by acute immune perturbations^12,14^. Despite substantial immune remodeling induced by cancer or its treatment, CMV reactivity persisted, suggesting a resilient memory T-cell compartment. Interestingly, our data showed no significant age-related decline in CMV responses, contrasting with some studies reporting reduced CMV-specific functionality in the elderly^9–11^. This discrepancy could be due to differences in sample sizes, but may also reflect our use of an integrated scoring method that captures residual functional activity better than spot counts alone.

One of the most notable findings from our study is the significant correlation between CMV-specific T-cell responses and the increase in spike-specific responses following vaccination, particularly in individuals without cancer and patients treated with immunotherapy. This suggests that CMV reactivity may serve as a marker of immunological "fitness," reflecting an individual’s capacity to respond to novel antigens. Our results support the idea that persistent CMV exposure may bolster heterologous immunity in some contexts, as previously reported for influenza^8,12,14^. Importantly, most of these studies dichotomized participants into CMV-seropositive and CMV-seronegative groups. This binary classification likely masks the underlying gradient of CMV-specific immunity, which may vary substantially even among seropositive individuals. By directly measuring CMV-specific T-cell responses and using continuous scoring, we were able to uncover associations that would be obscured by serostatus alone. Our findings, therefore, help reconcile conflicting reports by showing that it is not merely CMV exposure (i.e., seropositivity), but the magnitude of CMV-specific memory that predicts vaccine responsiveness.

Interestingly, we observed that this correlation between CMV and spike responses was weakened in patients treated with chemotherapy, suggesting that chemotherapy-induced alterations to the T-cell pool may disrupt the beneficial effects of CMV memory. Recent studies have shown that systemic therapies, particularly those used in hematologic malignancies and solid tumors, can induce long-lasting disruptions to T-cell compartments, including loss of naive and memory T-cell diversity, changes in TCR clonality, and a shift toward dysfunctional or exhausted states. For example, long-term survivors of multiple myeloma exhibit sustained immune alterations, including skewed T-cell phenotypes and reduced repertoire diversity, even decades after completing first-line therapy^24^. Similarly, acquired resistance to targeted therapies in melanoma has been shown to create an immune-evasive tumor microenvironment that confers cross-resistance to immunotherapy, reflecting broader dysfunction in immune cell engagement and persistence^25^. These findings underscore the need to interpret CMV’s immunomodulatory effects within the context of cancer therapy exposure, as therapeutic history may shape not only the composition but also the functional potential of the T-cell pool. In line with this, our coefficient of variation analysis revealed that chemotherapy-treated cohorts exhibited the greatest heterogeneity in vaccine-induced spike responses. In contrast, immunotherapy-treated patients and controls without cancer showed more consistent responses. Furthermore, individuals with lower CMV-specific responses tended to have higher variability in spike responses, while those with higher CMV responses showed more uniform outcomes. This relationship could indicate that stronger CMV memory stabilizes the magnitude of recall responses, or that individuals are inherently better at generating memory responses and perform well for both CMV and spike antigens.

Our study thus highlights the potential of CMV-specific T-cell reactivity as a biomarker of vaccine readiness, especially in immunocompromised populations. The association between CMV memory and heterologous responses may reflect shared survival niches, cross-reactivity, or low-level stimulation of innate immune pathways via persistent inflammation^8,13^. However, it may also signal immune over-activation or exhaustion in some contexts. Thus, future studies should assess not just the quantity but the quality and phenotype of CMV-specific T-cells, such as their cytokine production, expression of exhaustion markers, and subset composition (e.g., CD8+ TEMRA cells).

Importantly, our approach provides a scalable framework for integrating image-based immunometrics into large immunological studies. The open-source nature of *CMVision* enables broader application across vaccine trials, aging studies, and immune monitoring programs. Given that ELISpot is widely used in clinical immunology, improving its accuracy and resolution can have significant translational value.

## Data Availability

The original contributions presented in the study are included in the article. Further inquiries can be directed to the corresponding author. The CMVision pipeline and related code generated in this work can be found at https://github.com/d-bhatt/CMVision.

## Ethics statement

The study involving humans was approved by Declaration of Helsinki, Good Clinical Practice guidelines (ClinicalTrials.gov, NCT04715438)^18^. The study was conducted in accordance with the local legislation and institutional requirements. The human samples used in this study were acquired from primarily isolated materials. Written informed consent for participation was required and obtained for all participants and the trial protocol was approved by the medical ethics committee of the University Medical Centre Groningen.

## Acknowledgments

We thank the patients and their partners, as well as the medical staff, clinical trial staff, pharmacists, nurses, and technicians at the participating sites, the referring colleagues, VOICE consortium members, and the Department of Clinical Trials Office at the Netherlands Comprehensive Cancer Organization for their participation and support.

## Author contribution

**DKB**: Conceptualization, Writing and editing manuscript, Data collection and analysis, Software, Visualization, Funding acquisition

**MJ:** Conceptualization, Writing and editing manuscript, Data analysis, Software, Visualization

**FV:** Writing and editing manuscript, Data collection and analysis, Validation, Visualization

**AB:** Writing and editing manuscript, Data collection and analysis, Validation, Visualization

**SFO:** contributed to writing and editing the manuscript, study organization, data analysis, and funding.

**AAMvdV:** contributed to writing and editing the manuscript, study organization, data analysis, and funding.

**TJNH:** contributed to writing and editing the manuscript, study organization, data analysis, and funding.

**CHGvK:** contributed to writing and editing the manuscript, study organization, data analysis, and funding.

**A-MCD:** contributed to writing and editing the manuscript, study organization, data analysis, and funding.

**EFS:** contributed to writing and editing the manuscript, study organization, data analysis, and funding.

**JBAGH:** contributed to writing and editing the manuscript, study organization, data analysis, and funding.

**EGEdV:** contributed to writing and editing the manuscript, study organization, data analysis, and funding.

**DvB:** Conceptualization, Writing and editing manuscript, Supervision, Funding acquisition

